# Child and Adolescent Psychiatrists’ Use, Attitudes, and Understanding of Genetic Tests in Clinical Practice

**DOI:** 10.1101/2023.01.24.23284953

**Authors:** Takahiro Soda, Amanda R. Merner, Brent J. Small, Laura N. Torgerson, Katrina Muñoz, Jehannine Austin, Eric A. Storch, Stacey Pereira, Gabriel Lázaro-Muñoz

## Abstract

**Objective:** To report current practices and attitudes of child and adolescent psychiatrists (CAP) regarding diagnostic genetic and pharmacogenetic (PGx) testing.

**Methods:** Survey of 958 US-based practicing CAP.

**Results:** 54.9% of respondents indicated that they had ordered/referred for a genetic test in the past 12 months. 87% of respondents agreed that it is their role to discuss genetic information regarding psychiatric conditions with their patients; however, 45% rated their knowledge of genetic testing practice guidelines as poor/very poor. The most ordered test was PGx (32.2%), followed by chromosomal microarray (23.0%). 73.4% reported that PGx is at least slightly useful in child and adolescent psychiatry. Most (62.8%) were asked by a patient/family to order PGx in the past 12 months and 41.7% reported they would order PGx in response to a family request. Those who ordered a PGx test were more likely to have been asked by a patient/family and to work in private practice. 13.8% of respondents agreed/strongly agreed that a PGx test can predict the effectiveness of specific antidepressants. Some respondents also indicated they would make clinical changes based on PGx information even if a medication was currently effective and there were no side effects.

**Conclusions:** Genetic testing has become routine clinical care in child and adolescent psychiatry. Despite this, many providers rate their associated knowledge as poor/very poor. Patient requests were associated with ordering practices and providers misinterpretation of PGx may be leading to unnecessary changes in clinical management. There is need for further education and support for clinicians.

## Introduction

The past decade has seen the identification of hundreds of genomic loci associated with psychiatric conditions, the ability to generate polygenic risk scores for psychiatric conditions, and the emergence of commercially available pharmacogenetic (PGx) tests aimed specifically at psychotropic medications ^1–5^. A study of US-based psychiatrists found that 14% ordered genetic tests in the past 6 months, and 41% had patients ask about genetic testing^6^. A decade later, there are few data on current practices and attitudes towards genetic testing among adult or child and adolescent psychiatrists (CAP).

Genetic testing is currently part of the standard of care in the evaluation of children and adults for some neuropsychiatric conditions, including autism spectrum disorder (ASD)^7–9^ and intellectual and developmental disorders (IDD)^9,10^, where results can impact subsequent medical decision making^9,11,12^; (e.g. detecting inborn errors of metabolism can lead to specific treatments^13^, and genetic diagnoses can trigger screening and follow up for associated medical conditions ^9,12^. Numerous professional organizations recommend genetic testing to evaluate ASD, but implementation remains low ^11,14,15^.

A different category of genetic test can be used to guide pharmacological treatments^16,17^.

The FDA has approved medication inserts with pharmacogenetically-determined medication dose recommendations for certain medications, including commonly prescribed psychotropic medications such as citalopram^18^ and aripiprazole^19^. Clinical guidelines regarding PGx tests have been released^17,20^, including for carbamazepine dosing and *HLA-B* genotype^21^, and selective serotonin reuptake inhibitors for individuals carrying certain CYP2D6 or CYP2C19 alleles^20,22^. Currently, various commercial PGx laboratory-developed tests with accompanying decision-support tools are available for patients with psychiatric disorders^23^. Individual hospital-based laboratories have also developed tests that test panels of genes deemed relevant (e.g., Mayo Clinic, University of Florida).

The degree to which PGx testing can help guide optimal pharmacological treatment remains debated. Studies meta-analyzing the results from commercial tests have shown improved rates of remission for some tests, not for others^23–25^ when these were utilized relative to not.

Limitations of these studies include the subpopulations in which the studies were conducted and the discordance between results based on the specific test utilized. Professional organizations that represent psychiatrists in the United States (American Psychiatric Association, American Academy of Child and Adolescent Psychiatrists, American Society of Geriatric Psychiatrists, International Society of Psychiatric Genetics^26^) have not endorsed the routine use of these tests to date. What is clear is that a nuanced understanding and discussion of this topic is necessary instead of general statements that this broad class of testing can or cannot benefit care.

Given a decade of quick advances in psychiatric genomics research and debates about the utility of PGx, it is important to identify and understand psychiatrists’ current practices, understanding, and attitudes regarding genetic testing. Here we report on the survey results regarding current practices, knowledge, and attitudes regarding genetic testing used as part of clinical care with a focus on PGx testing.

## Method

### Design and Setting

The Institutional Review Board of Baylor College of Medicine approved this study (protocol number H-46219). Informed consent was obtained from all participants.

Survey methods have been previously described.^27^ Briefly, a 47-question survey was developed to assess CAP current practice, knowledge, and perceptions toward genetic testing based on current literature with input from an expert panel consisting of CAP, psychologists, genetic counselors, bioethicists, lawyers, and an anthropologist using a modified Delphi method^28^. The survey included three sections: general (diagnostic) genetic testing, PGx testing, and polygenic risk scores. Survey participants were recruited from publicly available listservs, professional organizations, national conferences, and other professional meetings. Web searches were conducted to identify other publicly available contact information for CAP. The survey was administered in English, and was electronically distributed over a four-week period in June 2020 using Qualtrics©. Questions about the current use of general genetic testing and PGx testing were used to learn about CAP knowledge, experience, opinions on current and potential future utility, and concerns and appropriateness of PGx testing. For results on knowledge and perceptions of the utility of genetic testing in the evaluation of ASD, see Soda et al.^15^, and for results related to polygenic risk scores, see Pereira et al.^27^. The survey is found as supplementary file 1.

### Measures

The survey items were designed to ascertain clinicians’ knowledge about genetic testing in psychiatry, experiences with ordering genetic testing, perceived current and future utility of genetic testing, and clinicians’ views on the potential impact of genetic testing on mental health stigma.

#### Self-rated knowledge of genetic testing

Respondents self-rated their knowledge about genetic testing in psychiatry, genetic testing guidelines in psychiatry, and knowledge about how to integrate genetic test results and PGx test results into practice. Response options included: Very Poor, Poor, Good, or Very Good.

#### Experiences/Current practices with ordering genetic testing

Ordering practices were ascertained using up to six questions nested in conditional response queues. The first question asked whether a respondent ordered any genetic testing in the past 12 months as a yes/no question. If the respondent selected yes, they were then asked the approximate percentage of patients for whom they ordered genetic tests and whether they involved a genetic specialist when considering and/or interpreting genetic tests.

Participants reported the conditions for which genetic tests were ordered (e.g., ASD, Obsessive Compulsive Disorder), the type of test ordered (e.g., Chromosomal microarray, targeted testing for a specific disorder), and their reasons for ordering (e.g., diagnostic clarification, medication side effects). All questions were multiple-choice with multiple selections enabled, and included an option to write in responses not listed.

#### Current and future utility of genetic testing in psychiatry

Respondents reported perceived current and future utility for genetic testing for ASD, IDD, for reasons other than for ASD/IDD, and child and adolescent psychiatry broadly.

Response options included: Not at all useful, Slightly useful, Useful, Very useful.

#### Attitudes toward role in genetic testing in psychiatry

Respondents reported whether they feel it is their role to discuss genetic information regarding psychiatric illness with patients and their families. Response options included: Strongly disagree, disagree, agree, and strongly agree.

#### Interpretation and clinical translation of knowledge of pharmacogenetics

Respondents reported whether there is sufficient evidence that PGx test results can predict the effectiveness of one antidepressant medication over another on a 5-point scale ranging from 1 (“Strongly disagree”) to 5 (“Strongly agree”). Respondents were also allowed to answer, “I do not know”. Respondents were also asked, “If PGx test results show that a patient is at an increased risk for a serious side effect, but the patient has responded well to the medication without any significant side effects, how likely are you to:” change the medication or change the dose of the current medication. Response options included: Very unlikely, Unlikely, Likely, or Very likely.

### Statistical Methods

Categorical variables were analyzed using chi-square and logistic regression. When cell sizes were small, Fisher’s exact Test was used to compare the categorical groups. For comparisons of the current or future utility of genetic testing, McNemar changes tests^29^ were computed. Continuous variables were assessed using analysis of variance and two-sided p-values (*p* < .05) are reported.

## Results

The survey was distributed to 5,677 individuals. Of 1180 who agreed to participate, 962 completed surveys were obtained (16.9% completion rate), which reflects ∼11.6% of practicing U.S. CAP. Four participants were excluded because they did not indicate whether they ordered genetic testing in the past 12 months, for a final total of 958 respondents included in analyses. See Table 1 for participant characteristics.

**Table 1.**
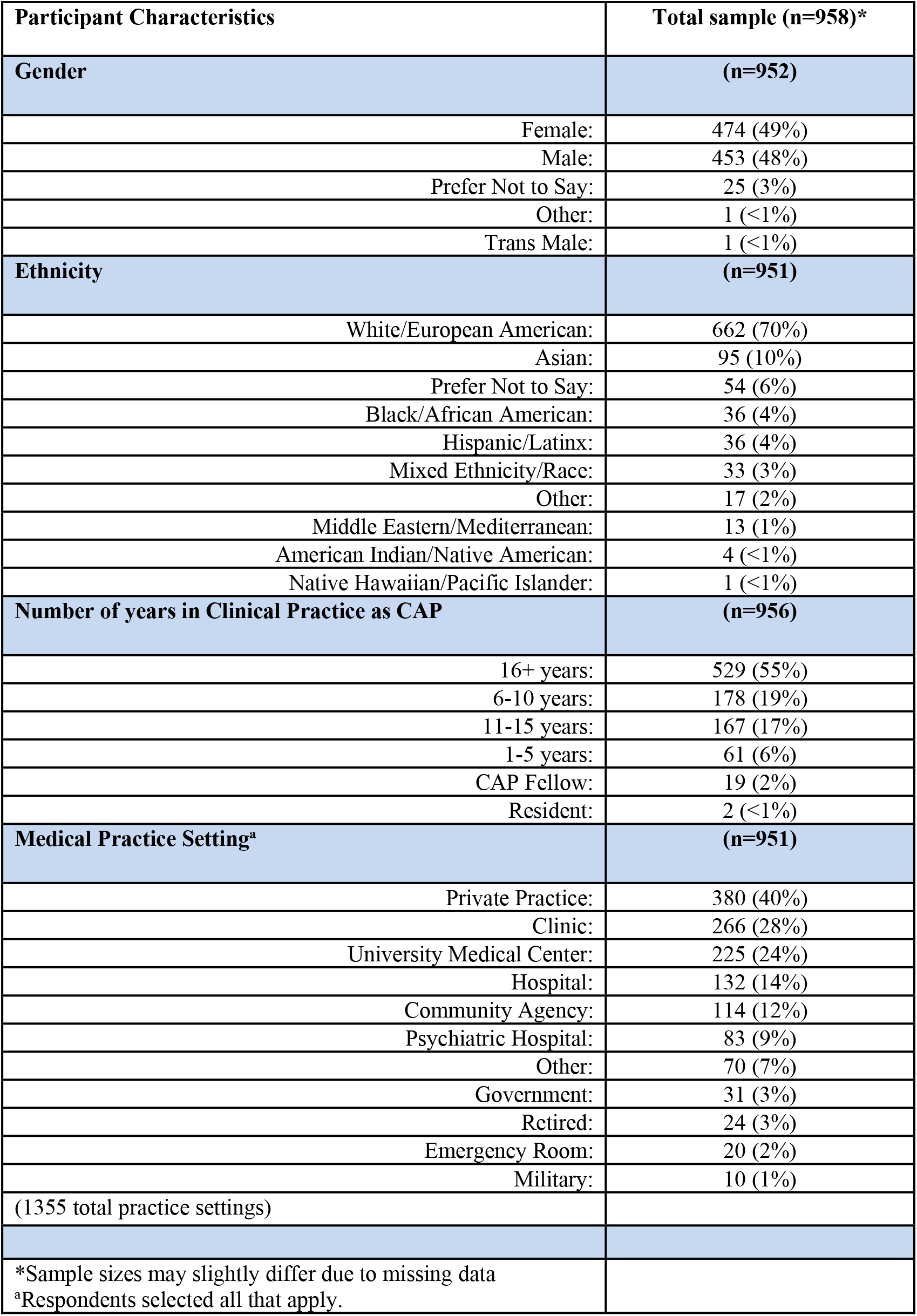
Demographic data

### Reported use and Subjective Knowledge of Genetic Testing

Fifty-five percent of respondents indicated they ordered/referred for any genetic test in the past 12 months, with the majority of those who ordered tests (86.3%) reporting ordering tests for ≤10% of their patients. Of those who ordered genetic testing, 60.5% reported that they involved a genetic specialist (e.g., a genetic counselor, medical geneticist, psychiatric geneticist).

Eighty-seven percent of respondents agreed that it is their role to discuss genetic information regarding psychiatric conditions with their patients and their families. However, 45.2% of all respondents, and 36.1% of those who ordered genetic tests, rated their subjective knowledge of genetic testing practice guidelines in psychiatry as poor or very poor. Furthermore, 33.2% of all respondents reported their knowledge of how to integrate genetic testing into their practice as poor or very poor, and 26.7% rated their knowledge of genetic testing in psychiatry as poor or very poor. However, with respect to PGx testing, 66.7% of respondents rated their knowledge of how to integrate PGx testing into their practice as good or very good.

The types of genetic tests ordered, conditions for which genetic testing was ordered, and reasons genetic testing was ordered by the CAP that responded as having ordered a genetic test in the past 12 months are shown in Figure 1. The most ordered test was PGx testing, followed by chromosomal microarray.

**Figure 1.**
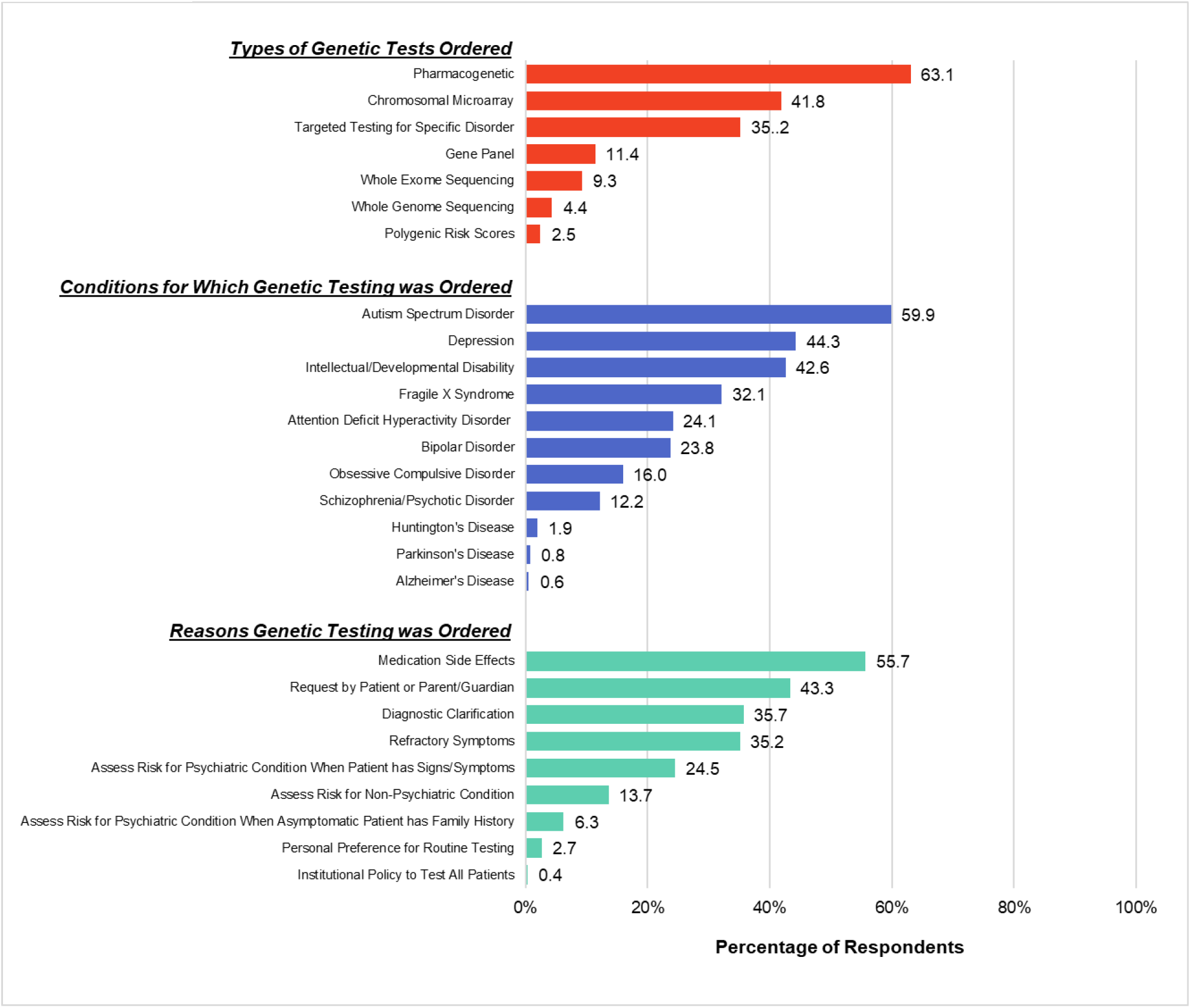
Respondents who ordered a genetic test in the past 12 months (n = 526) were asked what types of genetic tests they ordered, for which conditions they ordered the testing, and for which reasons they ordered the testing. The percentage of respondents choosing each option are shown. Please note that percentages will total over 100%, as respondents were able to select as many options that applied.

Regarding PGx testing, 32.2% of respondents reported ordering a PGx test in the prior 12 months. When asked when they would order a PGx test, the top three reasons were following refractory symptoms (55.0%), after severe side effects (50.3%), and when a family requested testing (41.4%; Figure 2). Fourteen percent of respondents reported they would never order PGx testing.

**Figure 2.**
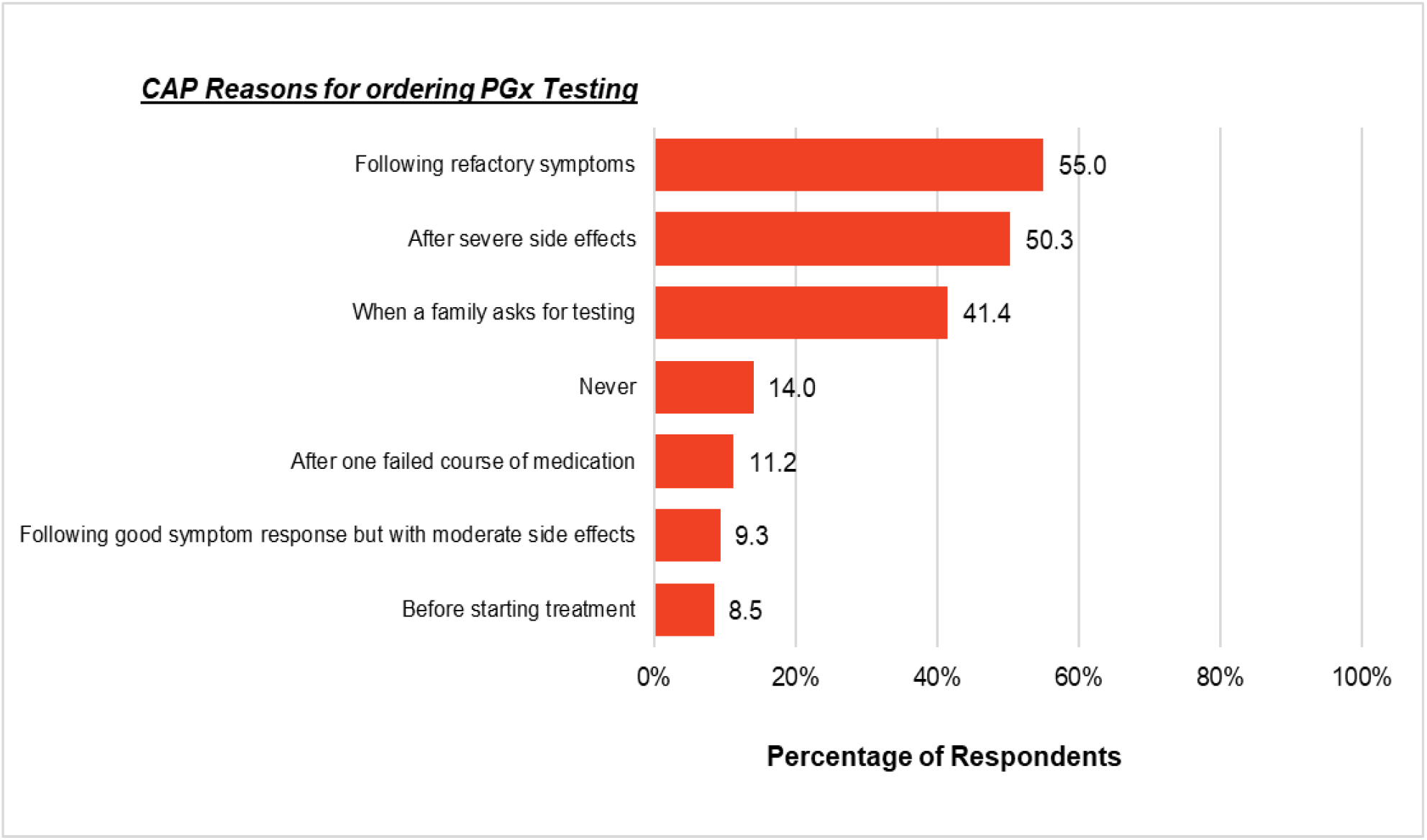
Percentage of respondents (n = 958) who chose each option when asked when they would order a PGx test. Please note that percentages will total over 100%, as respondents were able to select as many options that applied.

Given the only clinical indication for the use of CMA in psychiatry are for the medical evaluation of those with ASD/IDD, we examined whether respondents reporting that they ordered testing for ASD/IDD were generally the ones ordering CMA testing. Of the 958 respondents, 220 (23.0%) reported requesting a CMA. Of those who requested a CMA, 96.8% endorsed ordering tests for ASD or IDD in the past 12 months, as compared to 18.4% of providers ordering tests for ASD or IDD but reporting that they did not request a CMA in the past 12 months. This difference was statistically significant (OR = 134.69, 95% CI = 62.02 – 292.51, *p* <.001).

### Perceived utility

73.4% of respondents reported that PGx tests are *currently* at least slightly useful in CAP practice, and 93.3% believed PGx would be at least slightly useful in *five years* (X^2^(1) = 186.13, *p* <.001). We also identified a subgroup of respondents who reported ordering PGx testing despite identifying they felt PGx currently had no utility in child and adolescent psychiatry (7.1%). The most common reason these individuals reported ordering PGx tests was a family member requesting testing (68.2%).

In comparison, approximately 84% of respondents indicated that genetic testing was at least slightly useful for ASD (previously published in Soda et al.^15^, and 90.7% of respondents indicated that genetic testing was at least slightly useful for IDD.

Among CAP who ordered a genetic test in the previous 12 months, 87.7% rated genetic testing for ASD as slightly to very useful, whereas 82.1% of those who did not order a genetic test rated genetic testing for ASD as slightly to very useful (X^2^ (1) = 5.77, *p* =.017). Among CAP who ordered a genetic test, 93.2% rated genetic testing for IDD as slightly to very useful, whereas 89.0% of those who did not order a genetic test rated genetic testing as slightly to very useful (X^2^ (1) = 10.90, *p* <.001).

Overall, 76.3% of respondents felt that genetic testing for reasons other than testing for ASD/IDD was *currently* at least slightly useful. In contrast, 94.4% rated the utility of genetic testing for reasons other than ASD/IDD in *five years* would be slightly to very useful (X^2^(1) = 163.28, *p* <.001).

### Predictors of ordering a genetic test

Respondents’ perceived utility of genetic testing for conditions other than ASD/IDD was associated with an increased likelihood of ordering any genetic test (OR = 1.65, 95% CI = 1.21 – 2.24, *p* =.002). In addition to perceived utility, respondents were more likely to report ordering a genetic test when they reported greater knowledge about: genetic testing in psychiatry (OR = 2.20, 95% CI = 1.76 – 2.75, *p* <.001), genetic testing practice guidelines (OR = 1.83, 95% CI = 1.50 – 2.23, *p* <.001), and how to integrate genetic testing overall into their practice (OR = 2.35, 95% CI = 1.89 – 2.94, *p* <.001).

CAP with <11 years of post-fellowship years in practice were 37% less likely to request any genetic test compared to CAP with >11 years of post-fellowship years in practice (OR = 0.63, 95% CI = 0.47 – 0.85, *p* =.002).

Practice setting was associated with the frequency and self-reported knowledge with regard to genetic testing. Those who self-reported at least some component of having a private practice were more likely to order PGx tests compared to those who were not in any private practice (OR = 1.86, 95% CI = 1.41 – 2.45, *p* <.001). This subgroup was also less likely to order diagnostic tests (OR = 0.51, 95% CI = 0.38 – 0.69, *p* <.001). Those with at least some private practice setting did not significantly differ regarding self-reported knowledge about genetic testing F (1, 943) = 0.63, *p* =.427 or genetic testing practice guidelines in psychiatry F (1, 929) = 0.12, *p* =.725 compared to those who did not. In contrast, CAP at university medical centers were more likely to order diagnostic genetic testing defined as CMA, Fragile X, exome/genome, and other specific genetic tests (OR = 2.71, 95% CI = 1.97 – 3.73, *p* <.001). Practicing at a university medical center was also associated with greater self-reported knowledge of practice guidelines regarding psychiatric genetic testing F (1, 909) = 4.67, *p* = .031, and how to integrate them into practice F (1,924) = 5.10, *p* = .024.

Regarding PGx testing, respondents who reported higher levels of perceived utility of PGx were more likely to report ordering PGx testing in the last 12 months (OR = 1.78, 95% CI = 1.47 – 2.16, *p* <.001). 62.8% of respondents reporting that they had been asked by a patient or family member to order a PGx test in the last year. Of the 32.3% who reported ordering a PGx test in the prior year, those who reported ordering a PGx test were significantly more likely to have been asked by a patient or family member to order PGx testing (49.2%) compared to respondents who did not receive a patient or family request for testing (6.8%), X^2^ (1) = 172.86, *p* < .001.

### Understanding of PGx

Given the demonstrated common use of PGx in care at present, we further assessed the potential impact of PGx results on current patient care. Approximately 14% (13.8%) of respondents agreed or strongly agreed that a PGx test can predict the effectiveness of one antidepressant over another, indicating a problematic gap in knowledge concerning how to interpret PGx testing results, and 1.8% indicated that they did not know. Furthermore, 6.9% noted that they would likely or very likely change a medication, and 12.0% noted that they would change the dosage of the currently prescribed medication if PGx showed a patient is at an increased risk for serious side effects even if the patient has responded well to the medication without any significant side effects.

### Discussion

We report the largest survey of psychiatrists on the topic of genetic testing to date.

Relative to a prior study in 2011^6^, 1) Respondents continue to believe it is their role to discuss genetic findings in the context of psychiatric practice, 2) The fraction of psychiatrists reporting use of genetic testing has dramatically increased to a degree that it now constitutes a majority, and 3) A substantial fraction continue to report poor knowledge about these tests and how to incorporate them into practice.

Most respondents (54.9%) ordered a genetic test in the prior year, suggesting that genetic testing is now a common part of clinical care in child and adolescent psychiatry in the U.S. A 2011 survey found that 14% of U.S. psychiatrists ordered genetic tests^6^, suggesting a significant increase in this practice. More respondents reported ordering PGx testing than any other type of test (i.e., Fragile X, CMA, or WES), all of which currently have clear guidelines supporting their use in child and adolescent psychiatry^7–10^, a status that PGx testing does not currently have^30,31^. The most commonly reported type of PGx test were commercial combinatorial tests combined with clinical decision support tools for which the greatest evidence exists in adults with depression after a first failed medication^23,24,31^, a result that has not been consistently replicated in a non-industry sponsored study^32^ or in children and adolescents^33^. These combinatorial tests do include the specific genetic tests contained within some established guidelines, and their output often contradicts standard of care^33^.

Most respondents answered the questions about knowledge of PGx in a manner consistent with the strength of the evidence about the clinical utility of these tests in psychiatric practice. That 8% of respondents would change a currently effective medication that was not producing any side effects based on a PGx test result indicates that test usage may contribute to unnecessary and inappropriate medication changes.

In contrast to diagnostic testing in the context of ASD/IDD, wherein perceived utility was associated with the likelihood of test ordering, the perceived utility of PGx testing did not seem to influence clinicians’ reported ordering of PGx. CAP who reported ordering PGx testing were more likely to order these tests when the patients or families asked them to order this type of test or if they had a private practice location. This has implications for future attempts to influence the use of these tests; it may be the case that whereas clinician education may play a role in increasing the deployment of clinically indicated genetic testing for medical evaluation in the context of ASD/IDD, the same may not be the case for PGx testing. Leaving aside the issue of whether or not PGx testing has sufficient evidence significantly impacting clinical outcomes for its implementation in the context of psychiatry, which remains widely debated^3,16,17,25,30,31^, clinicians should be ordering tests that they believe will positively impact clinical care, and not simply because a patient or family asked for it. Educational interventions that guide CAP on how to talk to patients about their reasons for not ordering certain tests may be indicated, as has been done with the prescription of unnecessary antibiotics in the primary care context^34,35^. There may be factors other than patient requests to account for the association between private practice and increased likelihood of ordering PGx tests, such as fewer restrictions or less oversight from institution policies, on-site pharmacists, practice formularies, including third-party payers, towards PGx testing such that private-practice psychiatry may allow for CAP to order these tests at an increased frequency relative to other locations.

It should be noted that there are several scenarios in which it is clinically appropriate to order PGx tests. For example, the current FDA labeling for carbamazepine has a black box warning that states that patients with ancestry in genetically at-risk populations should be screened for the presence of HLA-B*1502 prior to initiating treatment with carbamazepine.

Patients testing positive for the allele should not be treated with carbamazepine unless the benefit clearly outweigh the risks^36^. There is concern that those respondents who answered that they would never order a PGx test are either unaware of such labeling or alternatively have decided that they would never prescribe carbamazepine.

The difference between the implementation of genetic testing in the medical evaluation of ASD/IDD and PGx that may account for the higher rates of PGx being ordered in the absence of psychiatric professional organization’s inclusion of PGx as a part of guideline-concordant care include more active direct patient-engagement by PGx laboratories that provide this testing^37^.

Companies that offer PGx have greatly facilitated such testing by lowering the barriers towards ordering PGx on the part of physicians.

Several limitations should be considered. First, despite being the largest survey about genetics among CAP to date, our response rate was modest. Second, CAP were limited to those in the U.S. and results may not generalize to CAP in other countries.

The association between higher self-rated knowledge and guideline-concordant diagnostic genetic testing in respondents with academic affiliations may reflect differences in the dissemination of guidelines and implementation of practice change between such institutions relative to non-academic affiliated providers. The lag time between the release of a clinical guideline and its translation into clinical practice is commonly quoted to take 17 years^38^. Given the relative recency of the AACAP practice parameters for ASD^7^ and IDD^39^, the increased likelihood of having ordered genetic testing amongst those <10 years out of training is not entirely surprising. Board recertification testing for CAP was once every 10 years, and many trained community psychiatrists may have never had the training experience to order or interpret diagnostic genetic tests in the context of psychiatry. What is clear from the results of this survey is that guidelines are insufficient, and more education, collaboration with genetics healthcare professionals (i.e., genetic counselors^40^), as well as resources to appropriately implement genetic testing in psychiatry, are required.

## Supporting information

Supplemental File 1-CAPG Survey

## Data Availability

De-identified survey data are available on request from the corresponding author. Data will be made available to researchers whose proposed use of the data has been approved by the study team.

## Acknowledgements

This research was supported by the National Institutes of Health under Award Numbers 3R00HG008689 and R01MH128676, and P50HD103555 for the use of the Clinical and Translational Core facilities.

## References

1. Lu H, Qiao J, Shao Z, Wang T, Huang S, Zeng P. A comprehensive gene-centric pleiotropic association analysis for 14 psychiatric disorders with GWAS summary statistics. BMC Med. 2021;19:314. doi:10.1186/s12916-021-02186-z

2. Brainstorm Consortium, Anttila V, Bulik-Sullivan B, et al. Analysis of shared heritability in common disorders of the brain. Science. 2018;360(6395):eaap8757. doi:10.1126/science.aap8757

3. Fan M, Bousman CA. Commercial Pharmacogenetic Tests in Psychiatry: Do they Facilitate the Implementation of Pharmacogenetic Dosing Guidelines? Pharmacopsychiatry. 2020;53(4):174–178. doi:10.1055/a-0863-4692

4. Murray GK, Lin T, Austin J, McGrath JJ, Hickie IB, Wray NR. Could Polygenic Risk Scores Be Useful in Psychiatry?: A Review. JAMA Psychiatry. 2021;78(2):210–219. doi:10.1001/jamapsychiatry.2020.3042

5. Müller DJ. Pharmacogenetics in Psychiatry. Pharmacopsychiatry. 2020;53(4):153–154. doi:10.1055/a-1212-1101

6. Salm M, Abbate K, Appelbaum P, et al. Use of genetic tests among neurologists and psychiatrists: knowledge, attitudes, behaviors, and needs for training. J Genet Couns. 2014;23(2):156–163. doi:10.1007/s10897-013-9624-0

7. Volkmar F, Siegel M, Woodbury-Smith M, King B, McCracken J, State M. Practice Parameter for the Assessment and Treatment of Children and Adolescents With Autism Spectrum Disorder. J Am Acad Child Adolesc Psychiatry. 2014;53(2):237–257. doi:10.1016/j.jaac.2013.10.013

8. Hyman SL, Levy SE, Myers SM, et al. Identification, Evaluation, and Management of Children With Autism Spectrum Disorder. Pediatrics. 2020;145(1):e20193447. doi:10.1542/peds.2019-3447

9. Manickam K, McClain MR, Demmer LA, et al. Exome and genome sequencing for pediatric patients with congenital anomalies or intellectual disability: an evidence-based clinical guideline of the American College of Medical Genetics and Genomics (ACMG). Genet Med Off J Am Coll Med Genet. 2021;23(11):2029–2037. doi:10.1038/s41436-021-01242-6

10. Siegel M, McGuire K, Veenstra-VanderWeele J, et al. Practice Parameter for the Assessment and Treatment of Psychiatric Disorders in Children and Adolescents With Intellectual Disability (Intellectual Developmental Disorder). J Am Acad Child Adolesc Psychiatry. 2020;59(4):468–496. doi:10.1016/j.jaac.2019.11.018

11. Harris HK, Sideridis GD, Barbaresi WJ, Harstad E. Pathogenic Yield of Genetic Testing in Autism Spectrum Disorder. Pediatrics. 2020;146(4):e20193211. doi:10.1542/peds.2019-3211

12. Stafford CF, Sanchez-Lara PA. Impact of Genetic and Genomic Testing on the Clinical Management of Patients with Autism Spectrum Disorder. Genes. 2022;13(4):585. doi:10.3390/genes13040585

13. Abdelhakim M, McMurray E, Syed AR, et al. DDIEM: drug database for inborn errors of metabolism. Orphanet J Rare Dis. 2020;15:146. doi:10.1186/s13023-020-01428-2

14. Moreno-De-Luca D, Kavanaugh BC, Best CR, Sheinkopf SJ, Phornphutkul C, Morrow EM. Clinical Genetic Testing in Autism Spectrum Disorder in a Large Community-Based Population Sample. JAMA Psychiatry. 2020;77(9):979–981. doi:10.1001/jamapsychiatry.2020.0950

15. Soda T, Pereira S, Small BJ, et al. Child and Adolescent Psychiatrists’ Perceptions of Utility and Self-rated Knowledge of Genetic Testing Predict Usage for Autism Spectrum Disorder. J Am Acad Child Adolesc Psychiatry. 2021;60(6):657–660. doi:10.1016/j.jaac.2021.01.022

16. Ramsey LB, Namerow LB, Bishop JR, et al. Thoughtful Clinical Use of Pharmacogenetics in Child and Adolescent Psychopharmacology. J Am Acad Child Adolesc Psychiatry. 2021;60(6):660–664. doi:10.1016/j.jaac.2020.08.006

17. Bousman CA, Bengesser SA, Aitchison KJ, et al. Review and Consensus on Pharmacogenomic Testing in Psychiatry. Pharmacopsychiatry. 2021;54(1):5–17. doi:10.1055/a-1288-1061

18. Annotation of FDA Label for citalopram and CYP2C19. PharmGKB. Accessed September 28, 2022. https://www.pharmgkb.org/labelAnnotation/PA166104852

19. Annotation of FDA Label for aripiprazole and CYP2D6. PharmGKB. Accessed September 28, 2022. https://www.pharmgkb.org/labelAnnotation/PA166104839

20. Hicks JK, Bishop JR, Sangkuhl K, et al. Clinical Pharmacogenetics Implementation Consortium (CPIC) Guideline for CYP2D6 and CYP2C19 Genotypes and Dosing of Selective Serotonin Reuptake Inhibitors. Clin Pharmacol Ther. 2015;98(2):127–134. doi:10.1002/cpt.147

21. Leckband SG, Kelsoe JR, Dunnenberger HM, et al. Clinical Pharmacogenetics Implementation Consortium guidelines for HLA-B genotype and carbamazepine dosing. Clin Pharmacol Ther. 2013;94(3):324–328. doi:10.1038/clpt.2013.103

22. Hicks JK, Sangkuhl K, Swen JJ, et al. Clinical pharmacogenetics implementation consortium guideline (CPIC) for CYP2D6 and CYP2C19 genotypes and dosing of tricyclic antidepressants: 2016 update. Clin Pharmacol Ther. 2017;102(1):37–44. doi:10.1002/cpt.597

23. Ontario Health (Quality). Multi-gene Pharmacogenomic Testing That Includes Decision-Support Tools to Guide Medication Selection for Major Depression: A Health Technology Assessment. Ont Health Technol Assess Ser. 2021;21(13):1–214.

24. Bousman CA, Arandjelovic K, Mancuso SG, Eyre HA, Dunlop BW. Pharmacogenetic tests and depressive symptom remission: a meta-analysis of randomized controlled trials. Pharmacogenomics. 2019;20(1):37–47. doi:10.2217/pgs-2018-0142

25. Brown LC, Stanton JD, Bharthi K, Maruf AA, Müller DJ, Bousman CA. Pharmacogenomic Testing and Depressive Symptom Remission: A Systematic Review and Meta-Analysis of Prospective, Controlled Clinical Trials. Clin Pharmacol Ther. 2022;112(6):1303–1317. doi:10.1002/cpt.2748

26. Genetic Testing Statement | ISPG - International Society of Psychiatric Genetics. Accessed November 29, 2022. https://ispg.net/genetic-testing-statement/

27. Pereira S, Muñoz KA, Small BJ, et al. Psychiatric polygenic risk scores: Child and adolescent psychiatrists’ knowledge, attitudes, and experiences. Am J Med Genet Part B Neuropsychiatr Genet Off Publ Int Soc Psychiatr Genet. 2022;189(7-8):293–302. doi:10.1002/ajmg.b.32912

28. Dalkey N, Helmer O. An Experimental Application of the DELPHI Method to the Use of Experts. Manag Sci. 1963;9(3):458–467. doi:10.1287/mnsc.9.3.458

29. McNEMAR Q. Note on the sampling error of the difference between correlated proportions or percentages. Psychometrika. 1947;12(2):153–157. doi:10.1007/BF02295996

30. AACAP. Clinical Use of Pharmacogenetic Tests in Prescribing Psychotropic Medications for Children and Adol. Accessed September 29, 2022. https://www.aacap.org/aacap/Policy_Statements/2020/Clinical-Use-Pharmacogenetic-Tests-Prescribing-Psychotropic-Medications-for-Children-Adolescents.aspx

31. Zubenko GS, Sommer BR, Cohen BM. On the Marketing and Use of Pharmacogenetic Tests for Psychiatric Treatment. JAMA Psychiatry. 2018;75(8):769–770. doi:10.1001/jamapsychiatry.2018.0834

32. Oslin DW, Lynch KG, Shih MC, et al. Effect of Pharmacogenomic Testing for Drug-Gene Interactions on Medication Selection and Remission of Symptoms in Major Depressive Disorder: The PRIME Care Randomized Clinical Trial. JAMA. 2022;328(2):151–161. doi:10.1001/jama.2022.9805

33. Vande Voort JL, Orth SS, Shekunov J, et al. A Randomized Controlled Trial of Combinatorial Pharmacogenetics Testing in Adolescent Depression. J Am Acad Child Adolesc Psychiatry. 2022;61(1):46–55. doi:10.1016/j.jaac.2021.03.011

34. Fleming-Dutra KE, Mangione-Smith R, Hicks LA. How to Prescribe Fewer Unnecessary Antibiotics: Talking Points That Work with Patients and Their Families. Am Fam Physician. 2016;94(3):200–202.

35. Altiner A, Berner R, Diener A, et al. Converting habits of antibiotic prescribing for respiratory tract infections in German primary care--the cluster-randomized controlled CHANGE-2 trial. BMC Fam Pract. 2012;13:124. doi:10.1186/1471-2296-13-124

36. Carbamazepine Extended-Release Capsules Rx Only. Accessed November 23, 2022. https://nctr-crs.fda.gov/fdalabel/services/spl/set-ids/ceb84aa4-2041-6e6e-e053-2995a90a0f24/spl-doc?hl=carbamazepine

37. Health C for D and R. Inova Genomics Laboratory - 577422 - 04/04/2019. Center for Devices and Radiological Health. Published February 19, 2020. Accessed November 29, 2022. https://wayback.archive-it.org/7993/20201219080850/ https://www.fda.gov/inspections-compliance-enforcement-and-criminal-investigations/warning-letters/inova-genomics-laboratory-577422-04042019

38. Morris ZS, Wooding S, Grant J. The answer is 17 years, what is the question: understanding time lags in translational research. J R Soc Med. 2011;104(12):510–520. doi:10.1258/jrsm.2011.110180

39. Siegel M, McGuire K, Veenstra-VanderWeele J, et al. Practice Parameter for the Assessment and Treatment of Psychiatric Disorders in Children and Adolescents With Intellectual Disability (Intellectual Developmental Disorder). J Am Acad Child Adolesc Psychiatry. 2020;59(4):468–496. doi:10.1016/j.jaac.2019.11.018

40. Austin J, Inglis A, Hadjipavlou G. Genetic Counseling for Common Psychiatric Disorders: An Opportunity for Interdisciplinary Collaboration. Am J Psychiatry. 2014;171(5):584–585. doi:10.1176/appi.ajp.2014.13101421

