## Supplemental File 1-CAPG Survey for "Child and Adolescent Psychiatrists’ Use, Attitudes, and Understanding of Genetic Tests in Clinical Practice"

Child & Adolescent Psychiatric Genomics Survey

Start of Block: Consent

Q1 Child & Adolescent Psychiatric Genomics Survey

 The purpose of this survey is to learn about **child and adolescent psychiatrists' knowledge, practices and attitudes of genetic testing.** This study is sponsored by the United States National Human Genome Research Institute of the National Institutes of Health (Grant #R00HG008689 05S1). 
  
  Your participation is voluntary and your responses will remain confidential. The survey will take approximately 10-12 minutes. You may skip any questions you do not want to answer. Clicking on the "I agree to participate" button below implies your consent. 
  
  Upon completion of the survey, you will have the opportunity to select a $10 gift card. To do this, you will need to provide an email address which will only be used for the purpose of providing you the gift card.
  
  Thank you for participating in and supporting this research. If you have any questions, please contact Dr. Gabriel Lázaro-Muñoz, Assistant Professor, Baylor College of Medicine, at or 713-798-2672, or Dr. Eric Storch, Professor of Psychiatry and Behavioral Sciences, Baylor College of Medicine, at or 713-798-3579.  You may also contact the Institutional Review Board (IRB) at 713-798-6970.

- I agree to participate. (4)
- I choose not to participate. (5)

End of Block: Consent

Start of Block: Knowledge

Display This Question:

If Child & Adolescent Psychiatric Genomics Survey The purpose of this survey is to learn about child... = I agree to participate.

Q2 How would you rate your **knowledge** about:

|  | Very poor (1) | Poor (2) | Good (3) | Very good (4) |
| --- | --- | --- | --- | --- |
| Genetic testing in psychiatry (1) |  |  |  |  |
| Genetic testing practice guidelines in psychiatry (2) |  |  |  |  |

Q3 How would you rate your **knowledge** about how to integrate the following into practice:

|  | Very poor (1) | Poor (2) | Good (3) | Very good (4) |
| --- | --- | --- | --- | --- |
| Genetic testing overall (1) |  |  |  |  |
| Pharmacogenetic (PGx) results (2) |  |  |  |  |
| Polygenic risk scores (PRS) (3) |  |  |  |  |

End of Block: Knowledge

Start of Block: Genetic Testing Utility

Q4 **In the past 12 months**, have you personally ordered **ANY** genetic test (e.g., chromosomal microarray, gene sequencing panel, whole exome/genome sequencing, PRS, PGx) in your practice?

- No (1)
- Yes (2)

Q5 In the **past 12 months**, have you ordered **ANY** genetic test **via another provider** (e.g., genetic counselor, medical geneticist, primary care provider, etc.)?

- No (1)
- Yes (2)

Display This Question:

If In the past 12 months, have you personally ordered ANY genetic test (e.g., chromosomal microarray... = Yes

Or In the past 12 months, have you ordered ANY genetic test via another provider (e.g., genetic... = Yes

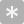

Q6 **In the past 12 months**, for approximately what ***percentage*** of patients have you ordered a genetic test personally or via another provider ?

________________________________________________________________

Display This Question:

If In the past 12 months, have you personally ordered ANY genetic test (e.g., chromosomal microarray... = Yes

Or In the past 12 months, have you ordered ANY genetic test via another provider (e.g., genetic... = Yes

Q7 **In the past 12 months**, when considering and/or interpreting genetic tests for your patients, have you involved a genetic specialist (e.g., genetic counselor, medical geneticist, psychiatric geneticist)?

- No (1)
- Yes (2)

Display This Question:

If In the past 12 months, have you personally ordered ANY genetic test (e.g., chromosomal microarray... = Yes

Or In the past 12 months, have you ordered ANY genetic test via another provider (e.g., genetic... = Yes

Q8 **In the past 12 months**, **for which conditions** have you ordered a genetic test personally or via another provider? (select all that apply)

- Autism Spectrum Disorder (ASD) (1)
- Intellectual / Developmental Disability (IDD) (2)
- Fragile X syndrome (3)
- Huntington’s disease (5)
- Alzheimer’s disease (7)
- Parkinson’s disease (8)
- Schizophrenia / psychotic disorder (9)
- Depression (10)
- Bipolar disorder (11)
- Attention Deficit Hyperactivity Disorder (ADHD) (14)
- Obsessive Compulsive Disorder (OCD) (12)
- Other: (please specify - e.g., breast cancer, sickle cell, coagulopathy, etc) (13) ________________________________________________

Display This Question:

If In the past 12 months, have you personally ordered ANY genetic test (e.g., chromosomal microarray... = Yes

Or In the past 12 months, have you ordered ANY genetic test via another provider (e.g., genetic... = Yes

Q9 **In the past 12 months,** **what type of genetic test** have you ordered personally or via another provider? (select all that apply)

- Chromosomal microarray (4)
- Whole exome sequencing (2)
- Gene panel (8)
- Whole genome sequencing (3)
- Pharmacogenetic (PGx) (1)
- Polygenic risk scores (PRS) (5)
- Targeted testing for a specific disorder (e.g., Fragile X syndrome) (6)
- Other: (please specify) (7) ________________________________________________

Display This Question:

If In the past 12 months, have you personally ordered ANY genetic test (e.g., chromosomal microarray... = Yes

Or In the past 12 months, have you ordered ANY genetic test via another provider (e.g., genetic... = Yes

Q10 **In the past 12 months**, **for which reasons** have you ordered a genetic test personally or via another provider? (select all that apply)

- Assess risk for a psychiatric condition when a patient has signs or symptoms of a disorder (3)
- Assess risk for a psychiatric condition when an asymptomatic patient has a family history of mental illness or ASD/IDD (4)
- Assess risk for a non-psychiatric condition (5)
- Diagnostic clarification (1)
- Medication side effects (6)
- Refractory symptoms (7)
- Request by patient or parent/guardian (9)
- Personal preference to routinely test my patients (2)
- Institutional policy is to order genetic testing for all patients (10)
- Other: (please specify) (11) ________________________________________________

End of Block: Genetic Testing Utility

Start of Block: Attitudes

Q11 **Currently,** **how useful** do you think genetic testing is for **Autism Spectrum Disorder (ASD)**?

- Not at all useful (1)
- Slightly useful (2)
- Moderately useful (3)
- Very useful (4)

Q12 Currently, **how useful** do you think genetic testing is for **Intellectual / Developmental Disability (IDD)**?

- Not at all useful (1)
- Slightly useful (2)
- Moderately useful (3)
- Very useful (4)

Q13 **Currently**, *other than for ASD/IDD,* **how useful** do you think genetic testing is in child and adolescent psychiatry?

- Not at all useful (1)
- Slightly useful (2)
- Moderately useful (3)
- Very useful (4)

Q14 **In 5 years**, *other than for ASD/IDD,* **how useful** do you think genetic testing will be in child and adolescent psychiatry?

- Not at all useful (1)
- Slightly useful (2)
- Moderately useful (3)
- Very useful (4)

Q15 I feel **it is my role to discuss** genetic information regarding psychiatric disorders with patients and their families.

- Strongly disagree (1)
- Disagree (2)
- Agree (3)
- Strongly agree (4)

End of Block: Attitudes

Start of Block: Pharmacogenetics

Q16 There is sufficient evidence to show that pharmacogenetic (PGx) results can **predict the effectiveness** of one antidepressant medication over another.

- Strongly disagree (1)
- Disagree (2)
- Neither agree nor disagree (3)
- Agree (4)
- Strongly agree (5)
- I don’t know (6)

Q17 If PGx test results show that a patient is at an increased risk for a serious side effect, *but* the patient has responded well to the medication without any significant side effects, **how likely are you to…**

|  | Very unlikely (1) | Unlikely (2) | Likely (3) | Very likely (4) |
| --- | --- | --- | --- | --- |
| Change medication (2) |  |  |  |  |
| Change dosage of current medication (3) |  |  |  |  |

Q18 **When** would you order a PGx test? (select all that apply)

- Before starting treatment (1)
- After **one** failed course of medication (2)
- Following good symptom response but with moderate side effects (3)
- When a family asks for testing (4)
- Following refractory symptoms (5)
- After severe side effects (6)
- Never (7)
- Other: (please specify) (8) ________________________________________________

Q19 In the past **12 months,** has a patient or their parent/guardian asked you to order a PGx test?

- No (1)
- Yes (2)

Q20 In the past **12 months,** have you ordered PGx tests personally or via another provider?

- No (1)
- Yes (18)

Display This Question:

If In the past 12 months, have you ordered PGx tests personally or via another provider? = Yes

Q21 **Which PGx tests** have you ordered? (select all that apply)

- Myriad - GeneSight psychotropic (1)
- Genomind (2)
- Other: (please specify) (3) ________________________________________________

Q22 **Currently, how useful** do you think PGx tests are in child and adolescent psychiatry?

- Not at all useful (1)
- Slightly useful (2)
- Moderately useful (3)
- Very useful (4)

Q23 **In** **5 years,** **how useful** do you think PGx tests will be in child and adolescent psychiatry?

- Not at all useful (1)
- Slightly useful (2)
- Moderately useful (3)
- Very useful (4)

End of Block: Pharmacogenetics

Start of Block: Polygenic Risk Scores

Q24 How would you **rate your knowledge** about polygenic risk scores (PRS)?

- I have never heard of PRS (1)
- Very poor (2)
- Poor (3)
- Good (4)
- Very good (5)

Skip To: End of Block If How would you rate your knowledge about polygenic risk scores (PRS)? = I have never heard of PRS

| Page Break |
| --- |

Q25 In the past **12 months**, have you requested or generated psychiatric PRS for a patient?

- No (1)
- Yes (2)

Display This Question:

If In the past 12 months, have you requested or generated psychiatric PRS for a patient? = Yes

Q26 **How** were those PRS generated? (select all that apply)

- Direct to consumer (1)
- Research study (2)
- Online tool (e.g., Impute.me) (3)
- I don't know (4)
- Other: (please specify) (5) ________________________________________________

Q27 In the past **12 months**, have you recommended testing for psychiatric PRS?

- No (1)
- Yes (2)

Q28 In the past **12 months**, has a patient or their parent/guardian asked you about psychiatric PRS?

- No (1)
- Yes (2)

Q29 In the past **12 months**, has a patient or their family brought you PRS that they obtained without your involvement?

- No (1)
- Yes (2)

Q30 Which of the following would prompt you to request or generate a patient's psychiatric PRS? (select all that apply)

- Assess risk for a psychiatric condition when a patient has signs or symptoms of a disorder (1)
- Assess risk for a psychiatric condition when an asymptomatic patient has a family history of mental illness or ASD/IDD (2)
- Assess risk for a non-psychiatric condition (3)
- Diagnostic clarification (4)
- Medication side effects (5)
- Refractory symptoms (7)
- Request by patient or parent/guardian (9)
- Personal preference to routinely test my patients (12)
- Institutional policy is to order genetic testing for all patients (10)
- Nothing would prompt me to request or generate a patient's PRS (13)
- Other: (please specify) (11) ________________________________________________

Q31 If a child or adolescent with no current psychiatric diagnosis had a **“high” (top 5%) PRS** for a psychiatric disorder, I would… (select all that apply)

- Have first degree relatives tested (1)
- Prescribe medications to help decrease risk (2)
- Recommend psychotherapy (3)
- Increase monitoring of symptoms (4)
- Refer to a genetic specialist (e.g., genetic counselor, medical geneticist) (5)
- Recommend lifestyle changes (6)
- Recommend modifying parenting styles or practices (7)
- Recommend modifying aspects of the child or adolescent's life to decrease stress (8)
- Evaluate patient for symptoms related to psychiatric disorders (11)
- Request consult from a genetic specialist (e.g., genetic counselor, medical geneticist) (12)
- Use online resources to learn more (13)
- Do nothing (9)
- Other: (please specify) (10) ________________________________________________

Q32 How **concerned** are you, if at all, that a "high" (top 5%) psychiatric PRS result will...

|  | Not at all concerned (1) | Slightly concerned (2) | Somewhat concerned (3) | Very concerned (4) |
| --- | --- | --- | --- | --- |
| Lead to over-treatment of children and adolescents with sub-threshold signs or symptoms of a disorder (1) |  |  |  |  |
| Lead to genetic discrimination (5) |  |  |  |  |
| Reduce parents' expectations for children (6) |  |  |  |  |
| Have a negative impact on a patient's emotional well-being (7) |  |  |  |  |
| Lead to misguided decisions regarding family planning (8) |  |  |  |  |

| Page Break |
| --- |

Q33 How **appropriate** would **PRS testing** be for the following groups?

|  | Very inappropriate (1) | Inappropriate (2) | Appropriate (3) | Very appropriate (4) |
| --- | --- | --- | --- | --- |
| Children or adolescents with sub-threshold signs or symptoms of a disorder (1) |  |  |  |  |
| First episode patients (2) |  |  |  |  |
| First degree relatives of patients diagnosed with a psychiatric disorder (3) |  |  |  |  |
| First degree relatives of an asymptomatic patient with high (top 5%) psychiatric PRS (4) |  |  |  |  |
| First degree relatives of patients diagnosed with a psychiatric disorder AND a high (top 5%) psychiatric PRS (5) |  |  |  |  |

Q34 How **appropriate** do you believe it is to **screen the general population** for psychiatric PRS?

- Very inappropriate (1)
- Inappropriate (2)
- Appropriate (3)
- Very appropriate (4)

Q35 How **appropriate** do you believe it is to do **preimplantation screening of embryos** for psychiatric PRS?

- Very inappropriate (1)
- Inappropriate (2)
- Appropriate (3)
- Very appropriate (4)

Q36 **Currently, how useful** do you think PRS are in child and adolescent psychiatry?

- Not at all useful (1)
- Slightly useful (2)
- Moderately useful (3)
- Very useful (4)

Q37 In **5 years,** **how useful** do you think PRS will be in child and adolescent psychiatry?

- Not at all useful (1)
- Slightly useful (2)
- Moderately useful (3)
- Very useful (4)

| Page Break |
| --- |

Q38 Please look at the following example of a genetic risk score from Impute.me.

 
 
  
 
**The result above shows that:**

|  | Agree (1) | Disagree (2) | Unsure (3) |
| --- | --- | --- | --- |
| There is a chance of about 0.25% for the person to develop this condition. (1) |  |  |  |
| The person has a slightly higher chance than the average person to develop the condition. (2) |  |  |  |
| The person will definitely develop the indicated condition. (3) |  |  |  |
| The person has a chance of just over 50% to develop the condition. (4) |  |  |  |
| The person has a chance of over 90% to develop the condition. (5) |  |  |  |

Q39 **What proportion of the overall risk** of developing a psychiatric disorder can be explained by current PRS?

- 0 - 20% (1)
- 20 - 40% (2)
- 40 - 60% (3)
- 60 - 80% (4)
- 80 - 100% (5)
- I don't know (7)

End of Block: Polygenic Risk Scores

Start of Block: Demographics

Q40 What is your **gender**? (select all that apply)

- Male (1)
- Female (2)
- Trans female/Trans woman (5)
- Trans male/Trans man (6)
- Genderqueer/Gender non-conforming (7)
- Different identity: (please specify) (3) ________________________________________________
- I prefer not to say (4)

Q41 What category or categories **best describe you**? (select all that apply)

- American Indian, Native American, Alaska Native (1)
- Asian (2)
- Black or African American (3)
- Hispanic/Latinx (8)
- Middle Eastern or Northern African/Mediterranean (7)
- Native Hawaiian/Pacific Islander (4)
- White or European American (5)
- Other: (please specify) (6) ________________________________________________
- I prefer not to say (9)

Q42 **How many years** have you been in clinical practice treating children and adolescents?

- General psychiatry resident (PGY 1-4) (1)
- Child and adolescent psychiatry fellow (2)
- 1-5 years post fellowship training (4)
- 6-10 years post fellowship training (5)
- 11-15 years post fellowship training (6)
- 16 or more years post fellowship training (10)
- I am not a child and adolescent psychiatrist. My speciality is: (please specify) (11) ________________________________________________

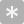

Q43 What **percentage of your patients** have Autism Spectrum Disorder **(ASD)** and/or Intellectual/ Developmental Disability **(IDD)**?

________________________________________________________________

Q44 What percentage of your patients fall within the following age groups?

< 12 years: : _______ (1)

12-17 years: : _______ (2)

> 18 years: : _______ (3)

Total : ________

Q45 Which of the following best describes your **practice setting**? (select all that apply)

- Private practice (1)
- Hospital (2)
- Clinic (6)
- Psychiatric hospital (3)
- Government (4)
- University medical center (5)
- Community agency (7)
- Military setting (8)
- Emergency room (9)
- Other: (please specify) (10) ________________________________________________

Q46 Would you consider participating in a **follow-up interview** to better understand psychiatrists' perspectives on genetic testing?  You would be compensated for your time.

- Yes, here is my email address: (2) ________________________________________________

Q47
If you would like a $10 gift card, please provide us with your **full name** and **email address** below so we can electronically send the gift card. 

Your responses will be kept confidential; your name and email will not be included in the data set.

- First name: (1) ________________________________________________
- Last name: (2) ________________________________________________
- Email: (3) ________________________________________________

End of Block: Demographics
